# The association of cardiovascular disease and other pre-existing comorbidities with COVID-19 mortality: A systematic review and meta-analysis

**DOI:** 10.1101/2020.05.10.20097253

**Authors:** Paddy Ssentongo, Anna E. Ssentongo, Emily S. Heilbrunn, Djibril M. Ba, Vernon M. Chinchilli

## Abstract

**Background:** Exploring the association of coronavirus-2019 disease (COVID-19) mortality with chronic pre-existing conditions may promote the importance of targeting these populations during this pandemic to optimize survival. The objective of this systematic review and meta-analysis is to explore the association of pre-existing conditions with COVID-19 mortality.

**Methods:** We searched MEDLINE, OVID databases, SCOPUS, and medrxiv.org for the period December 1, 2019, to May 1, 2020. The outcome of interest was the risk of COVID-19 mortality in patients with and without pre-existing conditions. Comorbidities explored were cardiovascular diseases (coronary artery disease, hypertension, cardiac arrhythmias, and congestive heart failure), chronic obstructive pulmonary disease, type 2 diabetes, cancer, chronic kidney disease, chronic liver disease, and stroke. Two independent reviewers extracted data and assessed the risk of bias. All analyses were performed using random-effects models and heterogeneity was quantified.

**Results:** Ten chronic conditions from 19 studies were included in the meta-analysis (n = 61,455 patients with COVID-19; mean age, 61 years; 57% male). Overall the between-study study heterogeneity was medium and studies had low publication bias and high quality. Coronary heart disease, hypertension, congestive heart failure, and cancer significantly increased the risk of mortality from COVID-19. The risk of mortality from COVID-19 in patients with coronary heart disease was 2.4 times as high as those without coronary heart disease (RR= 2.40, 95%CI=1.71-3.37, n=5) and twice as high in patients with hypertension as high as that compared to those without hypertension (RR=1.89, 95%CI= 1.58-2.27, n=9). Patients with cancer also were at twice the risk of mortality from COVID-19 compared to those without cancer (RR=1.93 95%CI 1.15-3.24, n=4), and those with congestive heart failure were at 2.5 times the risk of mortality compared to those without congestive heart failure (RR=2.66, 95%CI 1.58-4.48, n=3).

**Conclusions:** COVID-19 patients with all any cardiovascular disease, coronary heart disease, hypertension, congestive heart failure, and cancer have an increased risk of mortality. Tailored infection prevention and treatment strategies targeting this high-risk population are warranted to optimize survival.

## Introduction

The number of total cases of the coronavirus-2019 disease (COVID-19) continues to rise quickly, threatening thousands to millions of individuals with preexisting chronic conditions who are disproportionately affected.^1^ To date, May 5, 2020, the John Hopkins University coronavirus resource center reported that worldwide more than 180 countries have been affected with COVID-19 with more than three million confirmed cases and more than 250,000 deaths.^2^ As research related to potential risk factors for COVID-19 mortality continues, it is becoming clear that individuals with underlying conditions, such as cardiovascular disease (coronary artery disease, congestive heart failure [CHF], hypertension, and cardiac arrhythmias), cancer, chronic obstructive pulmonary disease (COPD), type 2 diabetes, chronic kidney disease (CKD), and chronic liver disease (CLD), all may be at an increased risk of death.^1,3,4^ As the number of published studies increase, there is a widening gap due to inconsistent findings related to the influence that types of pre-existing comorbidities have on COVID-19 mortality. Some studies report an association between preexisting conditions and COVID-19 mortality, whereas others report no association. With this being said, it is clear that regions experiencing the highest mortality rates, such as the United States, Europe, and China, also have the greatest burden of these preexisting chronic conditions.^5^

The novel virus, severe acute respiratory syndrome coronavirus 2 (SARS-CoV-2), the causative agent of COVID-19, interacts with angiotensin-converting enzyme 2 (ACE2), a cellular receptor expressed in the heart, kidney, pulmonary alveolar type II cells.^6^ It has been postulated, though not confirmed, that preexisting use of angiotensin II type 1 receptor blockers (ARBs) may upregulate membrane-bound ACE2 hence increasing susceptibility to virus entry.^7^ Therefore, it is plausible that individuals with preexisting chronic conditions such as high blood pressure and chronic heart failure taking ARBs may be more susceptible to the severity of SARS-CoV-2, including mortality.

To date, studies that have systematically explored the association of a range of pre-existing chronic conditions and COVID-19 mortality have limitation in the number of countries and number of conditions explored.^8,9^ We took a comprehensive approach and explored the association of major preexisting chronic conditions, including cardiovascular diseases such as coronary artery disease, CHF, stroke, hypertension, and cardiac arrhythmia, type 2 diabetes, COPD, asthma, cancer, HIV/AIDS, CKD, CLD, and stroke and the risk of mortality from COVID-19. Although the majority of studies occurred in China, we identified additional studies involving patients from Europe and North America. We hypothesize that the risk of mortality in COVID-19 patients is higher in patients with preexisting chronic conditions.

## Methods

We performed a systematic literature search of PubMed (MEDLINE), OVID (MEDLINE, HEALTHSTAR), SCOPUS, and medrxiv.org, using search criteria provided in the supplemental material (**Supplemental Document 1**). We followed the standards of the Meta-analysis of Observational Studies in Epidemiology (MOOSE, Supplemental Table 1).^10^ This initial search was supplemented by scanning of the references lists of relevant publications, and identifying their citations through the Web of Sciences (snowballing). We identified all studies published between October 1, 2019, and May 1st, 2020 reporting the risk of mortality in patients with COVID-19. Our search criteria included the following keywords and MesH terms: (“COVID-19” OR “Coronavirus” or “SARS-CoV-2”) AND (“Mortality”) AND (“cardiovascular disease” OR “chronic obstructive pulmonary disease” OR “asthma” OR “Coronary heart disease” OR “hypertension” OR “T2D” or “DM” or “diabetes” OR “cancer” OR “chronic kidney disease” OR “chronic liver disease” OR “comorbidities” OR “chronic disease”, OR” “HIV/AIDS” OR” Human immunodeficiency virus infection and acquired immune deficiency syndrome”). Two reviewers (ESH and AES) independently screened titles and abstracts of the studies for inclusion eligibility.

### Inclusion criteria

Studies were included in the analysis if they met the following inclusion criteria: COVID-19 was diagnosed based on the World Health Organization guidance;^11^ studies examined the association of any of the preexisting comorbidities and COVID-19; the risk point estimates reported as odds ratios (ORs), relative risks (RRs), or hazard ratios (HRs) or the data was presented such that the OR, (RR), or HR could be calculated; the 95% CI was reported, or the data were presented such that the 95% CI could be calculated. We excluded cases reports, studies not conducted on humans, review papers, meta-analyses, literature reviews and commentaries. All excluded studies were documented with reasons for their exclusion.

### Quality Assessment and Data Extraction

Since all of our studies were non-randomized observational studies, we used the Newcastle-Ottawa Scale (NOS) for quality assessment (Supplemental Document 2 and 3).^12^ Studies were first screened based on titles and abstracts by AES and ESH. If they met the inclusion criteria, then full-text was obtained and they were screened. In the event of disagreement, a third researcher (PS) was recruited to reach a consensus. Information extracted from included studies included title, year of publication, country, number of participants with each comorbidity who died and did not die, the RR/OR/HR of mortality of COVID-19 with each underlying condition, the mean or median age, proportion that was male, and other findings of interest.

### Data analysis

We adopted a narrative approach to describing the number of studies, study settings, diagnoses criteria for COVID-19, and the proportion of gender and race from each study. Our primary outcome was the risk of mortality in COVID-19 associated with preexisting chronic diseases. According to the previous study, if the outcome is rare in all populations and subgroups, the distinctions among different measures of RRs (e.g., odds ratios, rate ratios, and risk ratios) can be ignored ^13^, thus we combined RRs and HRs with ORs in the present meta-analysis and reported the pooled effect size as RRs as common risk estimates for all studies. We used the reported ORs, RRs, or HRs as the measures of the association between preexisting chronic conditions and the risk of mortality in COVID-19.

For studies without measures of associations, generalized linear mixed model were used to calculate the odds ratios using the number of events and the sample size of each study group.^14^ RRs, HRs and ORs in the present meta-analysis were reported as the pooled effect size as RRs as common risk estimates for all studies. First reported effect sizes were log-transformed to normalize the distributions. Second, standard errors (SEs) were calculated via the following equations ^15^: Lower = log (lower 95% CI) and upper = log (upper 95% CI), and SE = (upper - lower)/3.92. To assess the associations between preexisting conditions and the risk of mortality, we pooled the RR estimates for the presence versus absence of preexisting conditions from each study, weighted by the inverse of their variances (inter-study plus intra-study variances). The *metagen* function from the R package meta was used to calculate the pooled effect estimates using random-effects models.^16^ We invoked random-effects models to pool study results for the association between preexisting chronic conditions and the risk of mortality. Because of the few studies we identified, we did not conduct any meta-regression or subgroup analyses. ^16^ The DerSimonian and Laird (DL) random-effects method was used to estimate the pooled inter-study variance (heterogeneity).^17^ We graphically displayed individual and pooled estimates with forest plots. Inter-study heterogeneity was assessed using *I*^2^ statistics, expressed as % (low (25%), moderate (50%), and high (75%) and Cochrane’s *Q* statistic (significance level < 0.05) ^18,19^. Potential ascertainment bias (as might be caused by publication bias) was assessed with funnel plots, by plotting the study effect size against standard errors of the effect size, and Egger’s test.^20^ All statistical analyses with R software, version 3.4.3 (R, College Station, TX).

## Results

As shown in **Figure 1**, we identified a total of 261 studies from Scopus, OVID, PubMed, and Joana Briggs International EBF database. A total of 57 studies were excluded because as duplicates, leaving 204 studies to explore for inclusion. An additional 98 studies were excluded based on titles and abstracts and another 87 studies based on full text, which resulted in 19 studies for the quantitative analysis. The process yielded a total of 61,455 patients for the quantitative analysis, with 17 studies performed in China,^3,4,21-32^ 1 in Italy, and 1 that included 11 countries from North America, Europe, and Asia.^1^ (**Table 1**).

**Figure 1:**
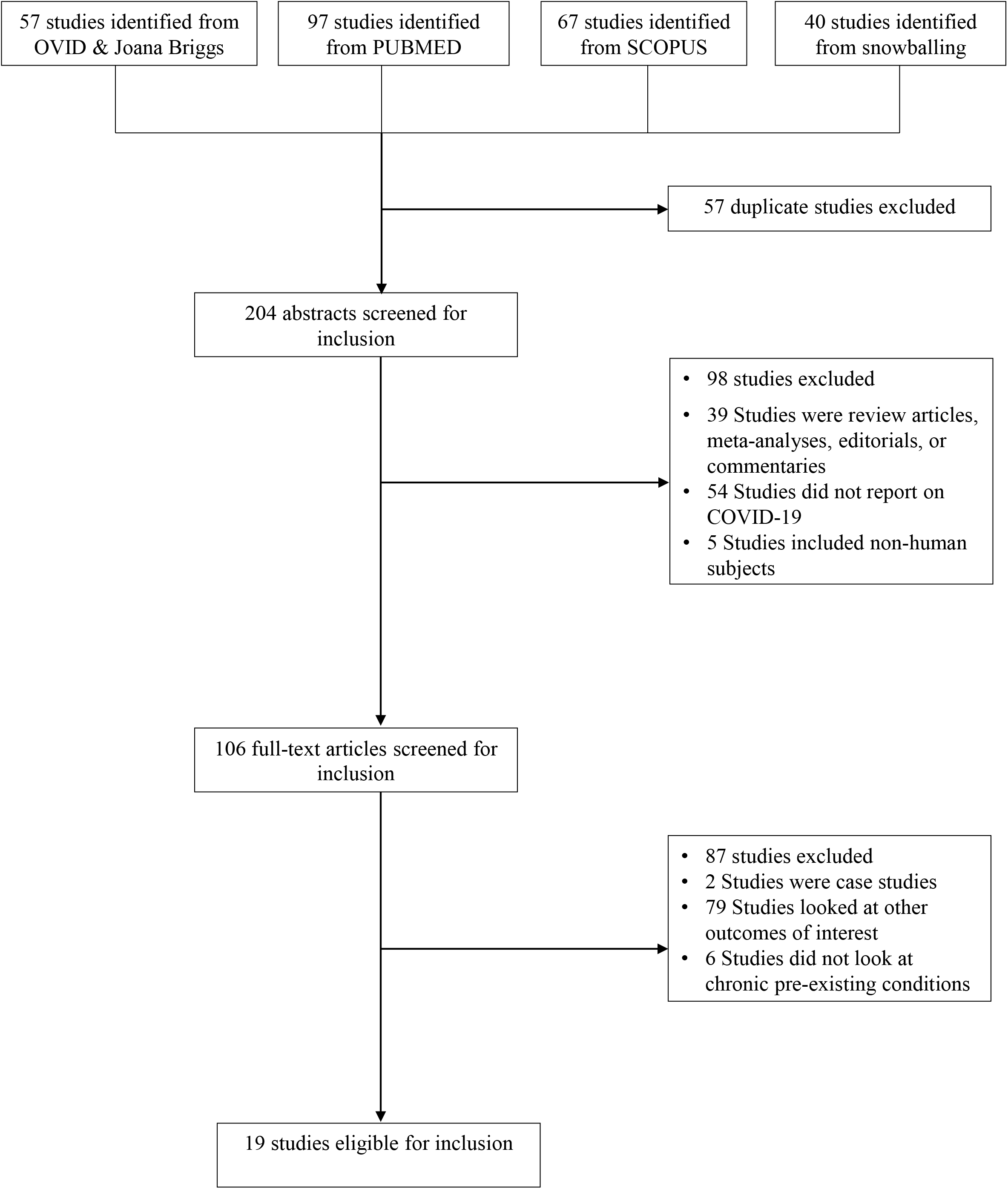
Flow Diagram

**Table 1:**
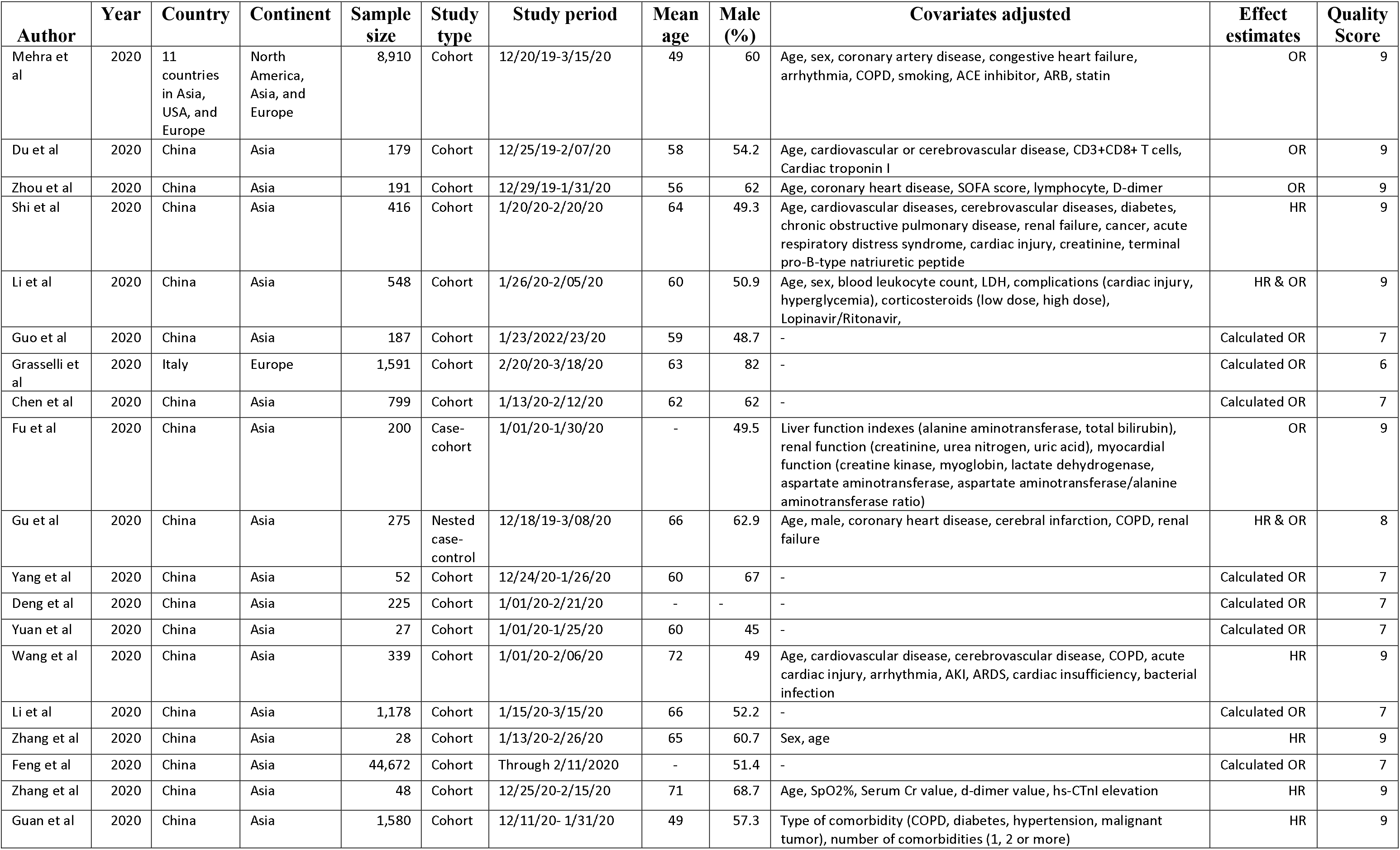
Characteristics of studies included in meta-analysis. Abbreviations: OR: Odds ratio; HR: Hazard ratio; COPD: Chronic obstructive pulmonary disease; ACE: Angiotensin-converting enzyme: ARB: Angiotensin II receptor blocker; hs-CTnI: high-sensitivity cardiac *troponin* I: AKI: Acute kidney injury; ARDS: Acute respiratory distress syndrome; SOAF: sequential organ failure assessment; LDH: Lactate dehydrogenase

| Author | Year | Country | Continent | Sample size | Study type | Study period | Mean age | Male (%) | Covariates adjusted | Effect estimates | Quality Score |
| --- | --- | --- | --- | --- | --- | --- | --- | --- | --- | --- | --- |
| Mehra et al | 2020 | 11 countries in Asia, USA, and Europe | North America, Asia, and Europe | 8,910 | Cohort | 12/20/19-3/15/20 | 49 | 60 | Age, sex, coronary artery disease, congestive heart failure, arrhythmia, COPD, smoking, ACE inhibitor, ARB, statin | OR | 9 |
| Du et al | 2020 | China | Asia | 179 | Cohort | 12/25/19-2/07/20 | 58 | 54.2 | Age, cardiovascular or cerebrovascular disease, CD3+CD8+ T cells, Cardiac troponin I | OR | 9 |
| Zhou et al | 2020 | China | Asia | 191 | Cohort | 12/29/19-1/31/20 | 56 | 62 | Age, coronary heart disease, SOFA score, lymphocyte, D-dimer | OR | 9 |
| Shi et al | 2020 | China | Asia | 416 | Cohort | 1/20/20-2/20/20 | 64 | 49.3 | Age, cardiovascular diseases, cerebrovascular diseases, diabetes, chronic obstructive pulmonary disease, renal failure, cancer, acute respiratory distress syndrome, cardiac injury, creatinine, terminal pro-B-type natriuretic peptide | HR | 9 |
| Li et al | 2020 | China | Asia | 548 | Cohort | 1/26/20-2/05/20 | 60 | 50.9 | Age, sex, blood leukocyte count, LDH, complications (cardiac injury, hyperglycemia), corticosteroids (low dose, high dose), Lopinavir/Ritonavir, | HR & OR | 9 |
| Guo et al | 2020 | China | Asia | 187 | Cohort | 1/23/2022/23/20 | 59 | 48.7 | - | Calculated OR | 7 |
| Grasselli et al | 2020 | Italy | Europe | 1,591 | Cohort | 2/20/20-3/18/20 | 63 | 82 | - | Calculated OR | 6 |
| Chen et al | 2020 | China | Asia | 799 | Cohort | 1/13/20-2/12/20 | 62 | 62 | - | Calculated OR | 7 |
| Fu et al | 2020 | China | Asia | 200 | Case-cohort | 1/01/20-1/30/20 | - | 49.5 | Liver function indexes (alanine aminotransferase, total bilirubin), renal function (creatinine, urea nitrogen, uric acid), myocardial function (creatinine kinase, myoglobin, lactate dehydrogenase, aspartate aminotransferase, aspartate aminotransferase/alanine aminotransferase ratio) | OR | 9 |
| Gu et al | 2020 | China | Asia | 275 | Nested case-control | 12/18/19-3/08/20 | 66 | 62.9 | Age, male, coronary heart disease, cerebral infarction, COPD, renal failure | HR & OR | 8 |
| Yang et al | 2020 | China | Asia | 52 | Cohort | 12/24/20-1/26/20 | 60 | 67 | - | Calculated OR | 7 |
| Deng et al | 2020 | China | Asia | 225 | Cohort | 1/01/20-2/21/20 | - | - | - | Calculated OR | 7 |
| Yuan et al | 2020 | China | Asia | 27 | Cohort | 1/01/20-1/25/20 | 60 | 45 | - | Calculated OR | 7 |
| Wang et al | 2020 | China | Asia | 339 | Cohort | 1/01/20-2/06/20 | 72 | 49 | Age, cardiovascular disease, cerebrovascular disease, COPD, acute cardiac injury, arrhythmia, AKI, ARDS, cardiac insufficiency, bacterial infection | HR | 9 |
| Li et al | 2020 | China | Asia | 1,178 | Cohort | 1/15/20-3/15/20 | 66 | 52.2 | - | Calculated OR | 7 |
| Zhang et al | 2020 | China | Asia | 28 | Cohort | 1/13/20-2/26/20 | 65 | 60.7 | Sex, age | HR | 9 |
| Feng et al | 2020 | China | Asia | 44,672 | Cohort | Through 2/11/2020 | - | 51.4 | - | Calculated OR | 7 |
| Zhang et al | 2020 | China | Asia | 48 | Cohort | 12/25/20-2/15/20 | 71 | 68.7 | Age, SpO2%, Serum Cr value, d-dimer value, hs-CTnI elevation | HR | 9 |
| Guan et al | 2020 | China | Asia | 1,580 | Cohort | 12/11/20- 1/31/20 | 49 | 57.3 | Type of comorbidity (COPD, diabetes, hypertension, malignant tumor), number of comorbidities (1, 2 or more) | HR | 9 |

### The risk of mortality from COVID-19 in patients with any cardiovascular disease

Of the nine studies that reported the risk of mortality associated with preexisting cardiovascular disease, 5 reported a significant association. The risk ratio point estimates for mortality from COVID-19 and cardiovascular disease ranged from 0.72 to 6.0 (**Figure 2A**). The overall pooled risk ratio of COVID-19 mortality associated with any cardiovascular disease was 2.35(95% CI: 1.44-3.84), implying a 2-fold increase in mortality. Between-study variation was moderate (I^2^= 65, p<0.01).

**2A.**
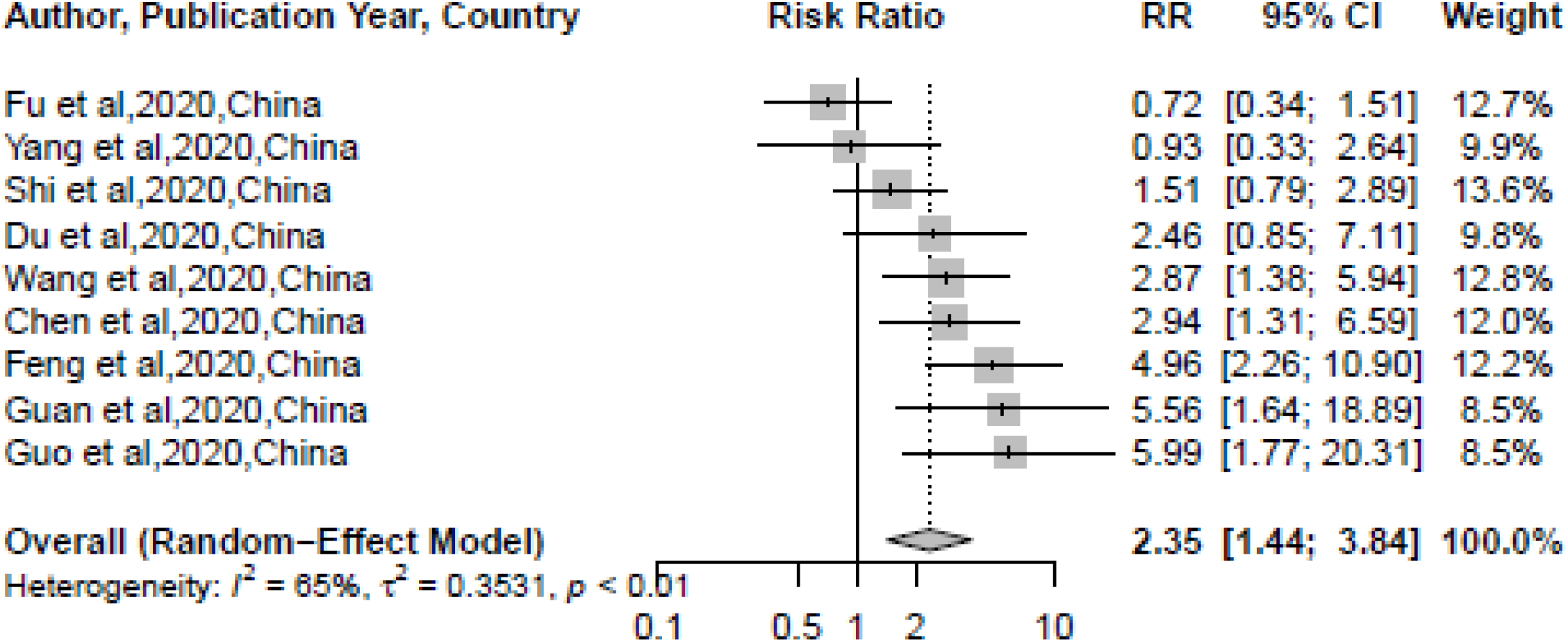
Cardiovascular disease

### The risk of mortality from COVID-19 in patients with preexisting chronic obstructive pulmonary disease

Three of seven studies that explored the association of preexisting COPD and COVID-19 mortality found that patients with COPD were at a significantly higher risk of mortality from COVID-19 compared to patients without COPD (**Figure 2B**). The pooled risk estimates for mortality from COVID-19 in patients with COPD was 1.76 (95% CI: 0.92-3.36), indicating that although the association was positive, it significant. Between-study heterogeneity was moderate (I^2^= 73, p<0.01).

**2B.**
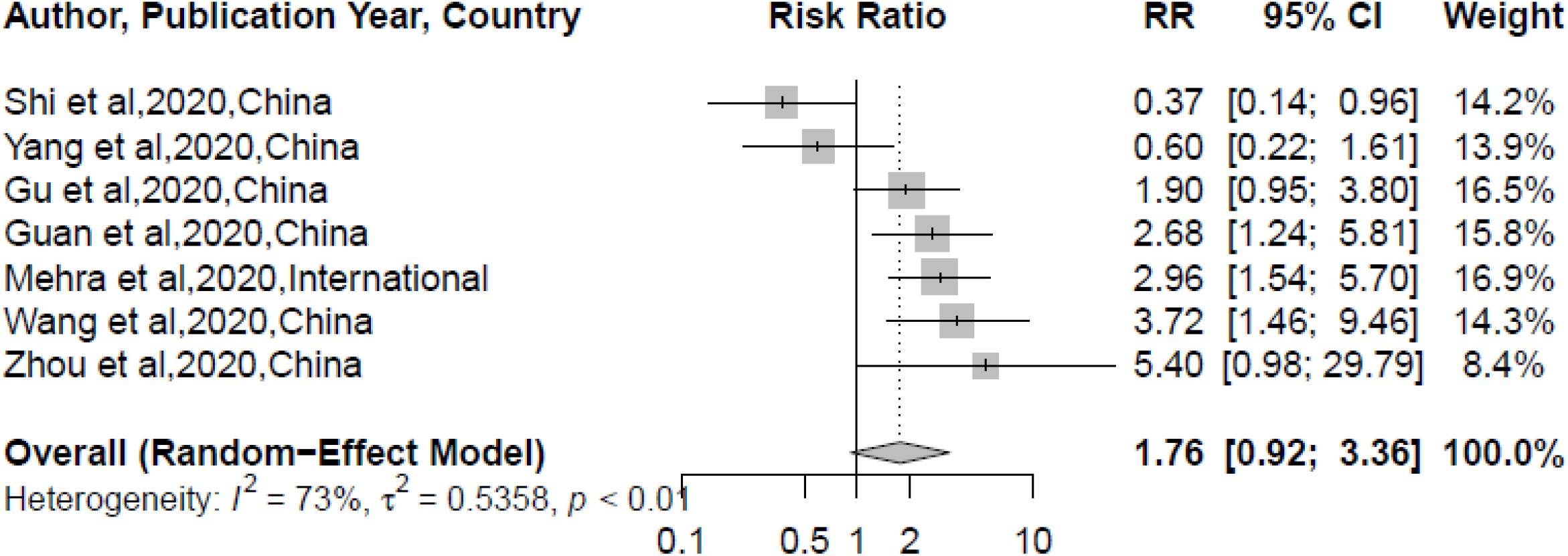
Chronic obstructive pulmonary disease

### The risk of mortality from COVID-19 in patients with preexisting coronary artery disease

Preexisting coronary artery disease significantly increased patient’s risk of mortality from COVID-19 (**Figure 2C**). Five studies reported the risk ratio of dying from COVID-19 with underlying coronary heart disease. Four studies were from China and one international study including patients from the United State. The risk ratio point estimates ranged from 1.3 to 3.2. Compared to COVID-19 patients without preexisting coronary heart disease, the risk of mortality in COVID-19 patients with coronary artery disease increased by 2-fold 95% CI: 1.58-2.27). The between-study heterogeneity was low (I^2^= 0%, p=0.79).

**2C.**
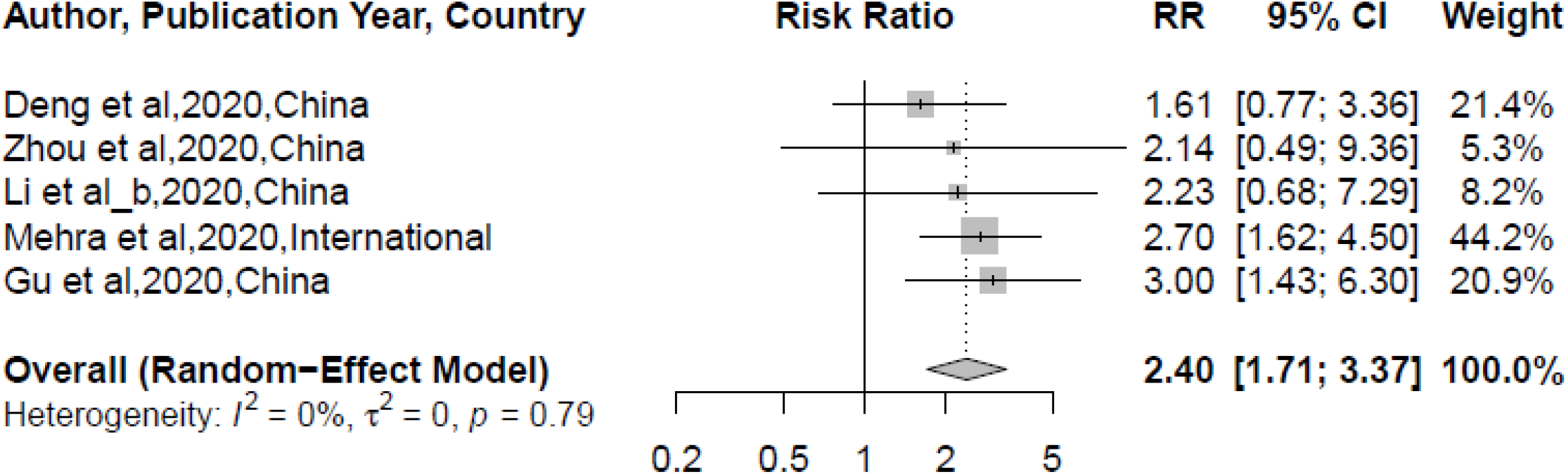
Coronary Heart Disease

### The risk of mortality from COVID-19 in patients with preexisting hypertension

Eight studies from China, and one from Italy, reported the risk of mortality from COVID-19 in patients with hypertension (**Figure 3A**). Patients with hypertension were nearly twice as likely to die from COVID-19 compared to patients without hypertension (RR: 1.89 95% CI: 1.58-2.27). Between-study heterogeneity was low (I^2^= 0%, p=0.61).

**3A.**
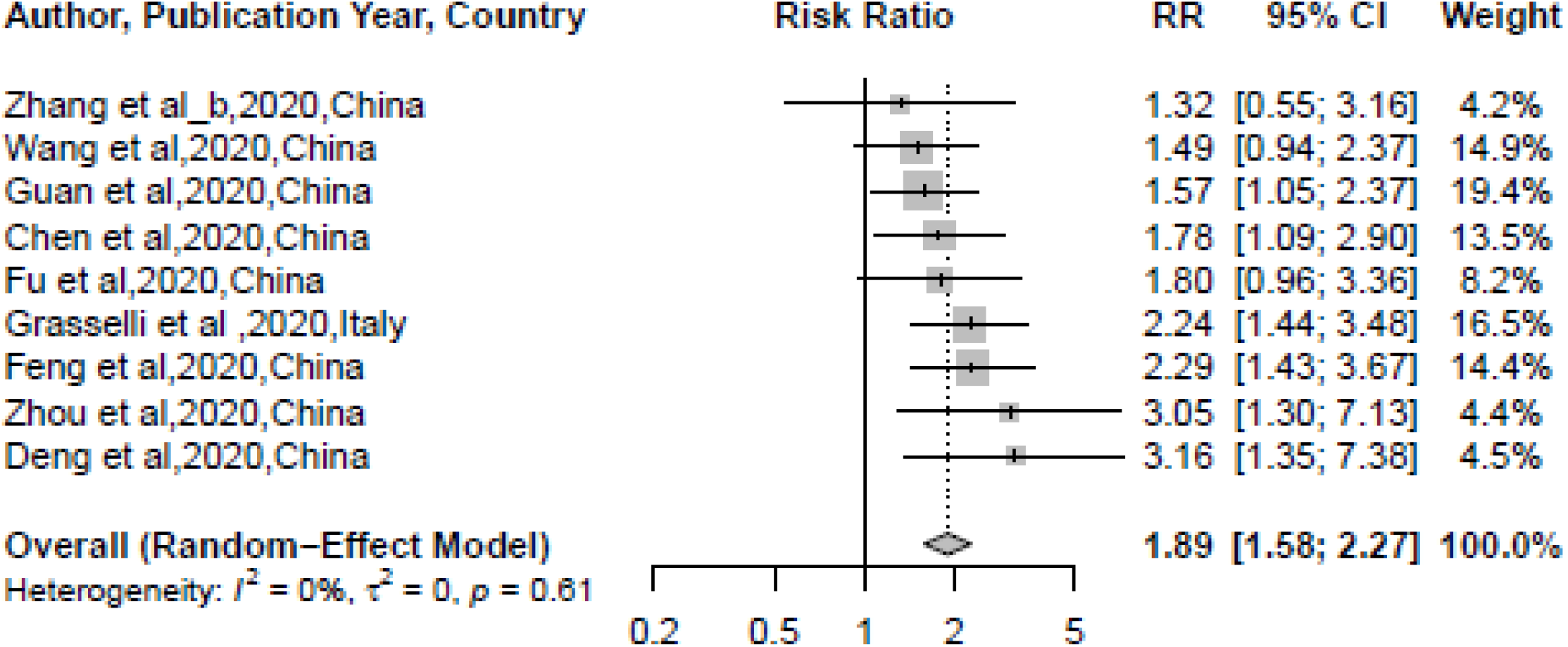
Hypertension

### The risk of mortality from COVID-19 in patients with preexisting type 2 diabetes

Twelve studies reported the risk of mortality from COVID-19 in patients with type 2 diabetes (**Figure 3B**). Of these, four studies found that patients with type 2 diabetes were at a significantly higher risk of mortality. Pooled estimates from random-effects models were not significant (RR:1.37, 95% CI: 0.85-2.20). Between-study heterogeneity was high (I^2^= 87%, p<0.01).

**3B.**
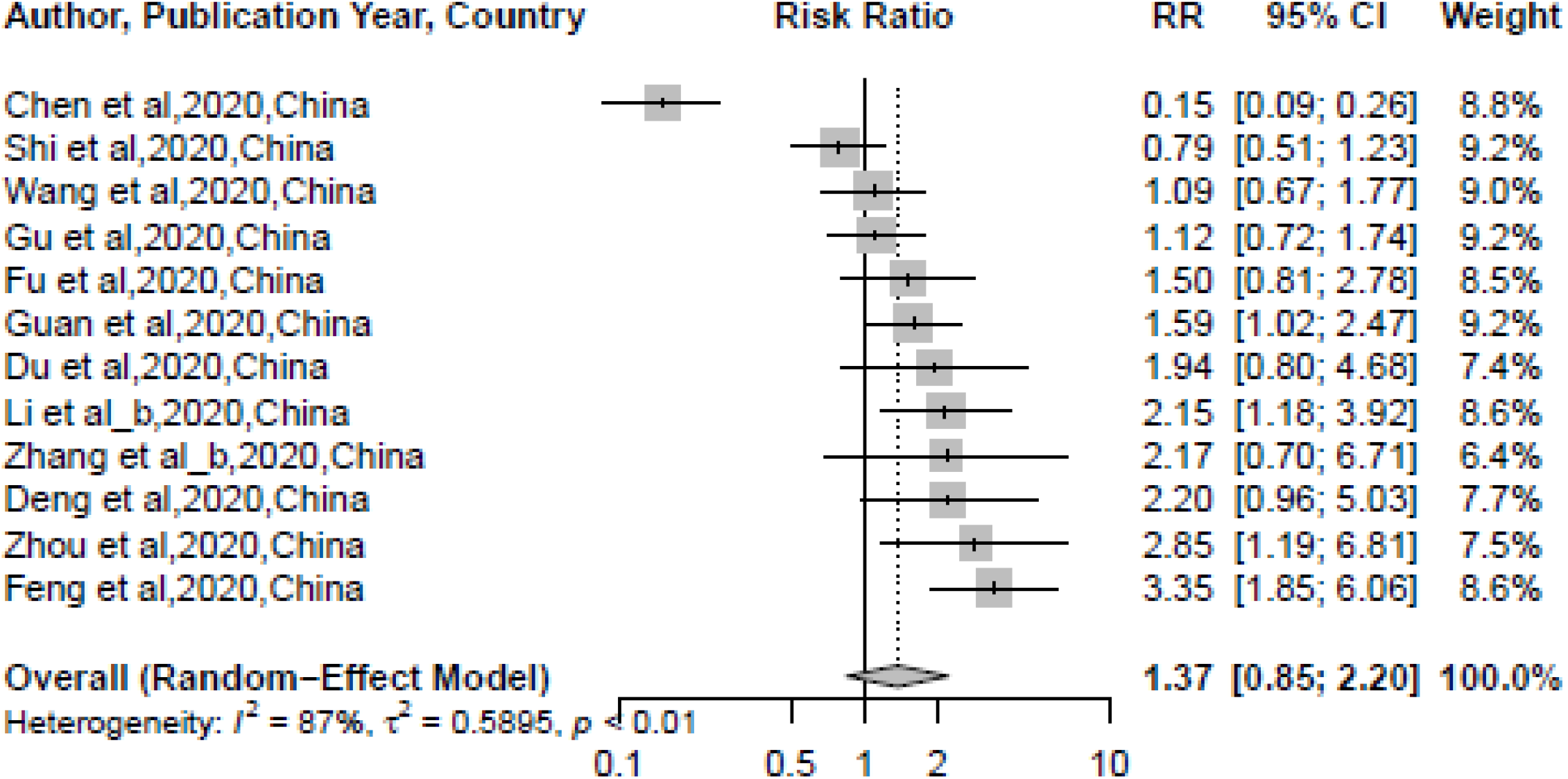
Type 2 Diabetes

### The risk of mortality from COVID-19 in patients with cancer

Four studies reported the COVID-19 mortality risk in patients with cancer (**Figure 3C**). Feng and colleagues and Guan and colleagues found that those with cancer were 2.5-3.5 times as likely to die from COVID-19 compared to those without cancer, however other studies found no significant association with COVID-19 mortality risk. Random-effects model pooled estimates was significant. COVID-19 patients with cancer were nearly twice as likely to die as COVID-19 patients without cancer (RR: 1.93, 95% CI: 1.15-3.24). Between-study heterogeneity was low (I^2^= 22%, p=0.28).

**3C.**
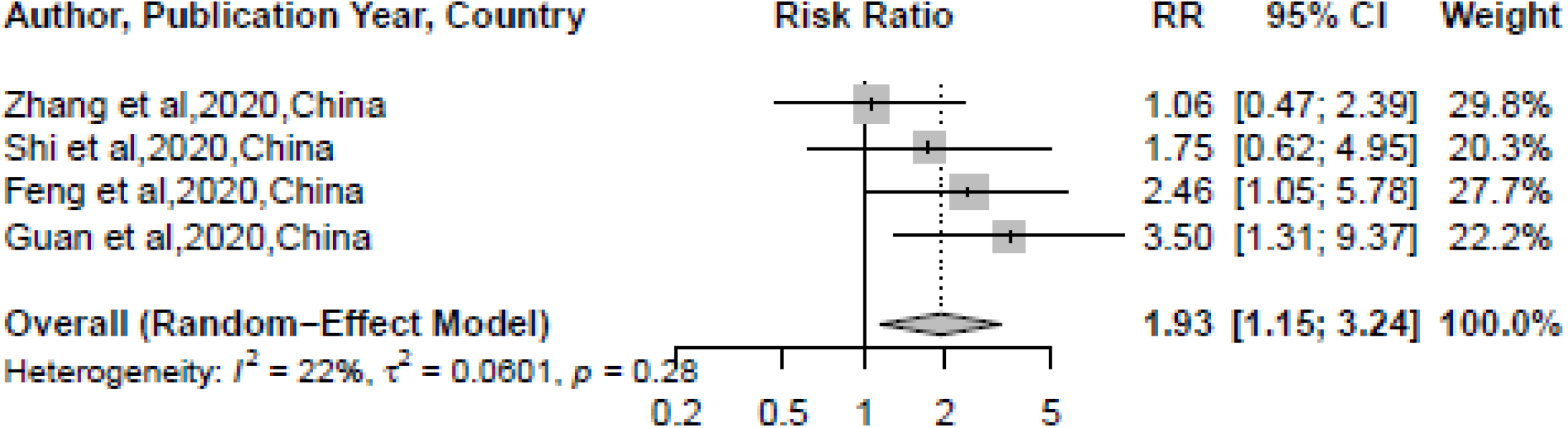
Cancer

### The risk of mortality from COVID-19 in patients with chronic kidney disease (CKD)

Four studies reported the risk of mortality from COVID-19 in patients with CKD (**Figure 4A**). One of these studies found that patients with chronic kidney disease were at increased risk of mortality from COVID-19. Pooled risk ratio estimates of 2,172 patients with CKD showed no significant association (RR: 2.36 95% CI: 0.97-5.77). Between-study heterogeneity was medium (I^2^= 63%, p=0.04).

**4A.**
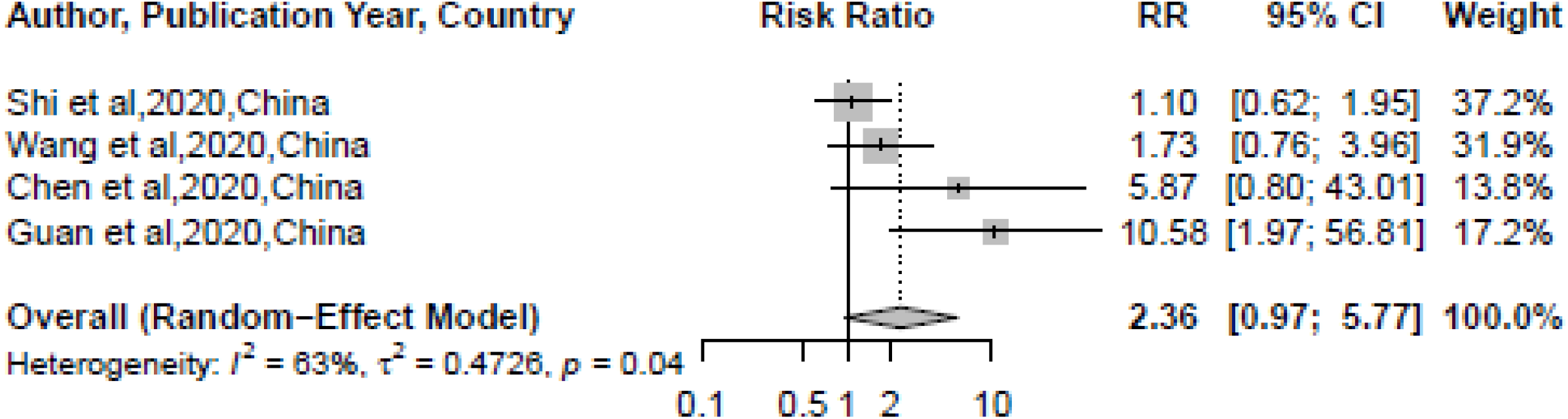
Chronic Kidney Disease

### The risk of mortality from COVID-19 in patients with chronic liver disease (CLD)

Two studies reported the association of CLD and risk of mortality from COVID-19 (**Figure 4B**). Both studies found no association CLD the risk of mortality. Pooled effect estimates were RR: 1.57 95% CI: 0.70-3.50. Between-study heterogeneity was low (I^2^= 0%, p=0.32).

**4B.**
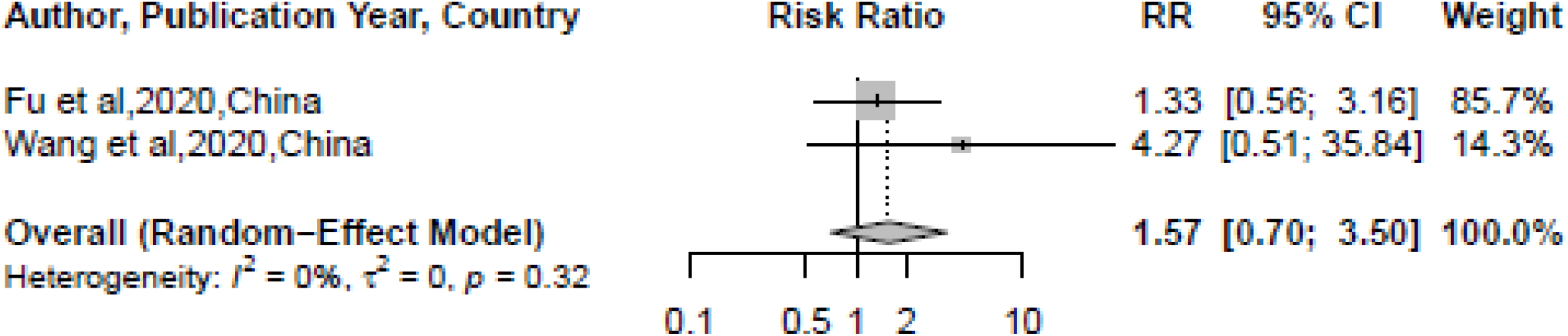
Chronic Liver Disease

### The risk of mortality from COVID-19 in patients with congestive heart failure (CHF)

Patients with CHF had a 3-fold higher risk of mortality from COVID-19 compared to those without CHF (**Figure 4C**, RR: 2.66, 95% CI: 1.58-4.48). Among the three studies reporting the risk of mortality from COVID-19 in patients with CHF, the risk ratio point estimates ranged from 2.44 to 3.89. Between-study heterogeneity low (I^2^= 0%, p=0.83).

**4C.**
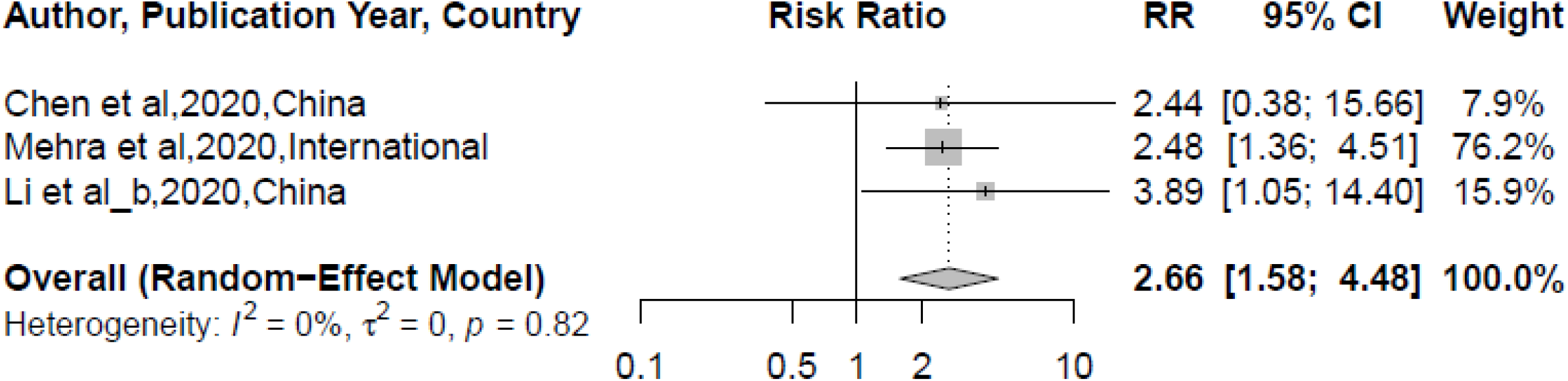
Congestive Heart Failure

### The risk of mortality from COVID-19 in patients with stroke

Three studies from China explored whether patients with a history of stroke were at an increased risk of COVID-19 mortality (**Figure 4D**). Two of the three studies found that patients with stroke were at a significantly higher risk of mortality, resulting in a pooled RR of 2.72, 95% CI: 0.90-8.21. Between-study heterogeneity high (I^2^= 75%, p=0.02).

**4D.**
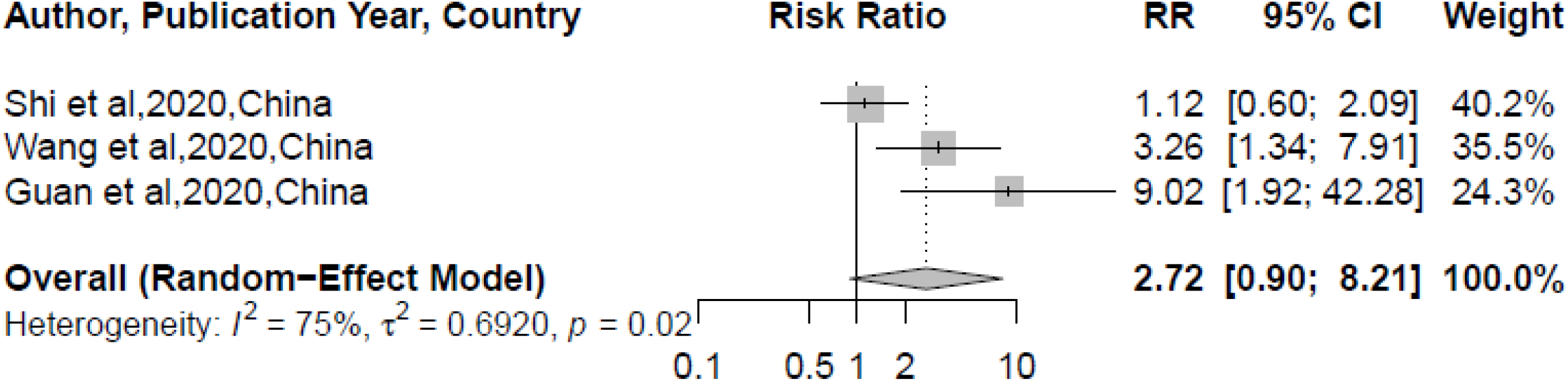
Stroke

One study reported the association between arrhythmia and COVID-19 mortality.^1^ COVID-19 patients with preexisting arrhythmia were 95% more likely to die compared to those without. OR: 1.95 (1.33– 2.86). We did not find studies reporting associations of HIV/AIDS and asthma and COVID-19 risk of mortality.

### Publication Bias and Study Quality

The funnel plot (Supplemental **Figure 1,2,3,4**) and the value of the Egger’s test (p=0.83,0.43.0.45.0.87 for figures 2A, 2B, 2C, 3A respectively) indicated absences of publication bias. The median study quality score for studies was 9 out of 9 (range=6–9, Table 1).

## Discussion

To the best of our knowledge, this is the first systematic review and meta-analysis to include COVID-19 cases from across Europe, Asia and North America, while also having the largest sample size (N=61,455) and number of studies (n=19). This meta-analysis was based on data from 19 studies with laboratory-confirmed COVID-19. Our findings suggest that patients with preexisting chronic conditions had a higher risk of death from COVID-19, particularly any cardiovascular disease, hypertension, coronary heart disease, CHF, and cancer. Our findings further highlight two important points. First, the failure of allostasis caused by preexisting conditions may in part explain the increased risk of mortality among COVID-19 patients. Second, a need for optimization of COVID-19 survival and limited health resources by employing focused vaccination for individuals with cardiovascular disease, CKD, and cancer.

A probable hypothesis of the pathophysiological mechanism related to the increased risk of mortality among COVID-19 patients may be explained by the allostatic load imposed on the body by cardiovascular and other preexisting conditions. Chronic conditions cause dysregulation of major physiological systems, including the hypothalamic-pituitary-adrenal axis, the sympathetic nervous system, and the immune system.^34^ The chronic nature of such conditions induces the ‘‘wear and tear’’ on the body’s regulatory systems,^35^ leading to the accumulation of pro-inflammatory cytokines, which affects the cellular immune system. As a result of the reduced immunity, these individuals become very susceptible to severe complications of SARS-CoV-2 and death. However, such an association with this type of virus is not relatively new. Seasonal influenza, SARS-CoV, and Middle Eastern respiratory syndrome (MERS)-CoV, have also been associated with increased severity and mortality in patients with preexisting conditions.^27,36^

As the race towards acquiring a vaccination against COVID-19 intensifies, the question remains as to which group of individuals should be prioritized for this vaccine. Our findings suggest that those with preexisting cardiovascular disease, hypertension, and cancer may benefit from vaccination to optimize both survival and limited resources. Targeted public health vaccination intervention strategy for influenza vaccination is recommended by the Advisory Committee on Immunization Practices against seasonal influenza.^37^ In the population with chronic comorbidities, annual influenza vaccination significantly reduces mortality and morbidity.^38^ Mounting evidence postulates that SARS-CoV-2 may become seasonal requiring annual vaccination.

Lastly, although the specific mechanisms are uncertain, SARS-CoV-2 is thought to infect host cells through ACE2 to cause COVID-19, while also causing damage to the myocardium.^39^ Majority of patients with preexisting cardiovascular disease use renin-angiotensin system (RAS) blockers, which are postulated to increase the risk of developing a severe and fatal SARSCoV-2 infection.^40^ In animal studies, Ferrario and colleagues reported ACE inhibitors or ARBs increased the levels of *Ace2* mRNA compared with placebo.^41^ Particularly, cardiac levels of *Ace2* mRNA increased by 4.7-fold or 2.8-fold with either lisinopril (an ACE inhibitor) or losartan (an ARB), respectively. However, whether or not patients with COVID-19 and pre-existing cardiovascular disease who are taking an ACE inhibitor or ARB should switch to another antihypertensive drug, remains controversial.^7^ Nevertheless, particular attention should be given to cardiovascular protection during treatment for COVID-19.

### Strengths and Limitations

As COVID-19 is still a relatively new phenomenon, there has been a limited number of comprehensive and conclusive studies related to mortality in diverse populations. Therefore, data from Africa and Australia were not included in this meta-analysis. We are hopeful that in the future, further research will be published that explores the association between COVID-19 mortality and preexisting chronic conditions in these regions. In addition to this, we were unable to explore the influence that cancer, HIV, and asthma may have on COVID-19 mortality. As mentioned above, further research is necessary to conclusively determine if individuals with these preexisting chronic conditions in these regions are at an increased risk for death from COVID-19.

As previously mentioned, this meta-analysis employed a large number of studies, which allowed for a relatively large sample size (n=61,455). By doing this, we were able to explore a broad scope of chronic conditions, with over seven comorbidities included. This allows our meta-analysis to comprehensively cover a multitude of prevalent conditions throughout populations.

## Conclusion

Our findings suggest that of the major comorbidities analyzed, coronary heart disease, hypertension, CHF, and cancer carry the strongest risk of death from COVID-19. This research highlights the importance of conducting further research related to the association between preexisting chronic conditions and COVID-19 mortality, in order to explore potential mechanisms to decrease this burden.

## Data Availability

All data is available within this manuscript.

## Notes

### Competing Interest Statement

The authors have declared no competing interest.

### Funding Statement

This study was not funded.

### Author Declarations

This is a meta-analysis and therefore it is IRB exempt.

